# Unveiling genetic architecture of white matter microstructure through unsupervised deep representation learning of fractional anisotropy maps

**DOI:** 10.1101/2025.07.04.25330856

**Authors:** Xingzhong Zhao, Ziqian Xie, Wei He, Hyun Yong Koh, Myriam Fornage, Degui Zhi

## Abstract

Fractional anisotropy (FA) derived from diffusion MRI is a widely used marker of white matter (WM) integrity. However, conventional FA-based genetic studies focus on phenotypes representing tract- or atlas-defined averages, which may oversimplify spatial patterns of WM integrity and thus limit the genetic discovery. Here, we proposed a deep learning–based framework, termed unsupervised deep representation of WM (UDR-WM), it adopted the voxel-wise FA maps as the input, and to extract brain-wide FA features—referred to as UDIP-FA—that capture distributed microstructural variation without prior anatomical assumptions. UDIP-FAs exhibit enhanced sensitivity to aging and substantially higher SNP-based heritability compared to traditional FA phenotypes (*P* < 2.20×10^-16^, Mann–Whitney U test, mean = 50.81%). Through multivariate GWAS, we identified 939 significant lead SNPs in 586 loci, mapped to 3480 genes, dubbed UDIP-FA related genes (UFAGs). UFAGs are overexpressed in glial cells, particularly in astrocytes and oligodendrocytes (*P <* 8.03× 10^-8^, Wald Test), and show strong overlap with risk gene sets for schizophrenia and Parkinson’s disease (P < 1.10 × 10^-4^, Fisher exact test). UDIP-FAs are genetically correlated with multiple brain disorders and cognitive traits, including fluid intelligence and reaction time, and are associated with polygenic risk for bone mineral density. Network analyses reveal that UFAGs form disease-enriched modules across protein–protein interaction and co-expression networks, implicating core pathways in myelination and axonal structure. Notably, several UFAGs, including *ACHE* and *ALDH2*, are targets of existing neuropsychiatric drugs. Together, our findings establish UDIP-FA as a biologically and clinically informative brain phenotype, enabling high-resolution dissection of WM genetic architecture and its genetic links to complex brain traits.

## Introduction

WM microstructure is a critical structural component of the brain that facilitate efficient communication between distributed gray matter regions, thereby supporting essential cognitive functions such as memory, attention, and executive control^1^.Fractional anisotropy (FA) is a widely used neuroimaging quantitative measure of WM microstructure, derived from the diffusion tensor images (DTI)^2^. FA is an important biomarker for studying brain function and brain-related disorders, as it sensitively captures WM microstructural changes associated with Alzheimer’s disease (AD) and various cognitive domains^3,4^, and several neurological and psychiatric disorders^5–8^. Understanding the genetic architecture of FA will inform the biological mechanisms of individual variability in cognitive and behavioral traits, as well as susceptibility to brain disorders.

Over the past decade, advances in imaging genetics have enabled large-scale genomic studies to elucidate the genetic architecture of FA. ENIGMA DTI Working Group and the Human Connectome Project demonstrated that FA derived phenotypes across major WM tracts exhibits moderate to high SNP-based heritability (*h²*: 0.53–0.90)^9^. Subsequent genome-wide association studies (GWAS) using UK Biobank imaging data identified genome-wide significant loci—such as *VCAN* (*rs67827860*)—linked to FA, suggesting shared genetic influences on both FA and WM hyperintensities^10^. A larger meta-GWAS combining the UK Biobank and CHARGE cohorts uncovered over 30 novel loci associated with FA, implicating genes involved in myelination and axonal integrity^11^. More recently, Zhao et al. analyzed diffusion MRI data from 43,802 individuals and identified 109 genomic regions related to WM microstructure—including FA—revealing a highly polygenic architecture with regulatory enrichment in oligodendrocytes and significant genetic correlations with numerous neuropsychiatric traits^12^. Additionally, a neonatal FA GWAS highlighted early developmental genetic effects, including an intronic SNP in *PSMF1*^9^.

Although these studies have revealed a part of the genetic component of FA, traditional FA GWAS face several methodological limitations that constrain their ability to fully capture the genetic architecture of WM microstructure. First, most studies rely on region- or tract-level mean FA intensity values based on predefined atlas, which substantially reduce the spatial complexity of the brain and limit the phenotypic variance explained. Second, traditional brain image GWAS frameworks typically assume linear and independent SNP effects on each phenotype dimension, which limits their ability to capture complex, nonlinear, and spatially distributed genetic influences across the brain. These limitations highlight the need for more integrative, high-dimensional approaches to advance our understanding of the genetic architecture of FA.

In this study, we developed and trained an unsupervised deep learning representation model that leverages whole-brain voxel-wise FA maps as model input to derive a set of global neuroimaging phenotypes, termed UDIP-FAs. Our model is based on the UDIP framework that was successfully applied to derive heritable phenotypes from T1- and T2-FLAIR-weighted images^13^. We then performed a multistage, multivariate GWAS of UDIP-FAs to delineate their genetic architecture. The UDIP-FA enabled us to elucidate the functional and biological mechanisms associated with FA and to investigate the associations between UDIP-FA and brain disorders at both the phenotypic and molecular levels. Our findings suggest that UDIP-FA serves as a robust global imaging phenotype, offering enhanced sensitivity for capturing the genetic architecture of WM and its links to neuropsychiatric and neurological disorders.

## Result

### An unsupervised representation framework to explore the genetic architecture of WM tracts using UDIP-FA

We developed a framework that extends unsupervised deep representation model to WM tracts (UDR-WM, Figure 1a, Figure S1). First, we pretrained an unsupervised 3D convolutional autoencoder on 6,130 FA maps derived from UK Biobank (UKB) diffusion tensor imaging (DTI) data (75% training, 25% validation; Methods and Figure 1a). Using this pre-trained UDR-WM, we extracted 128-dimensional deep learning representations—referred to as UDIP-FAs—from an independent UKB phase (N = 25,875). We found that these UDIP-FA dimensions were largely independent, with an average absolute pairwise Pearson correlation of 0.082 ± 0.065 (Figure S2). This suggests that each UDIP-FA dimension captures a distinct aspect of WM tract structure or variation. We then developed a suite of methods to interpret and apply these representations (Figure 1b) to investigate their underlying genetic architecture (Figure 1c). Finally, we systematically examined links between UDIP-FA features and brain disorders and related traits by integrating multi-omic and pharmacologic layers (genetic variants, genes, proteins, pathways, drug–target networks) with clinical phenotypes. (Figure 1d). In summary, our framework demonstrates that UDIP-FAs are informative and genetically meaningful markers for capturing the genetic architecture of WM tracts and for uncovering diverse disease associations.

**Figure 1:**
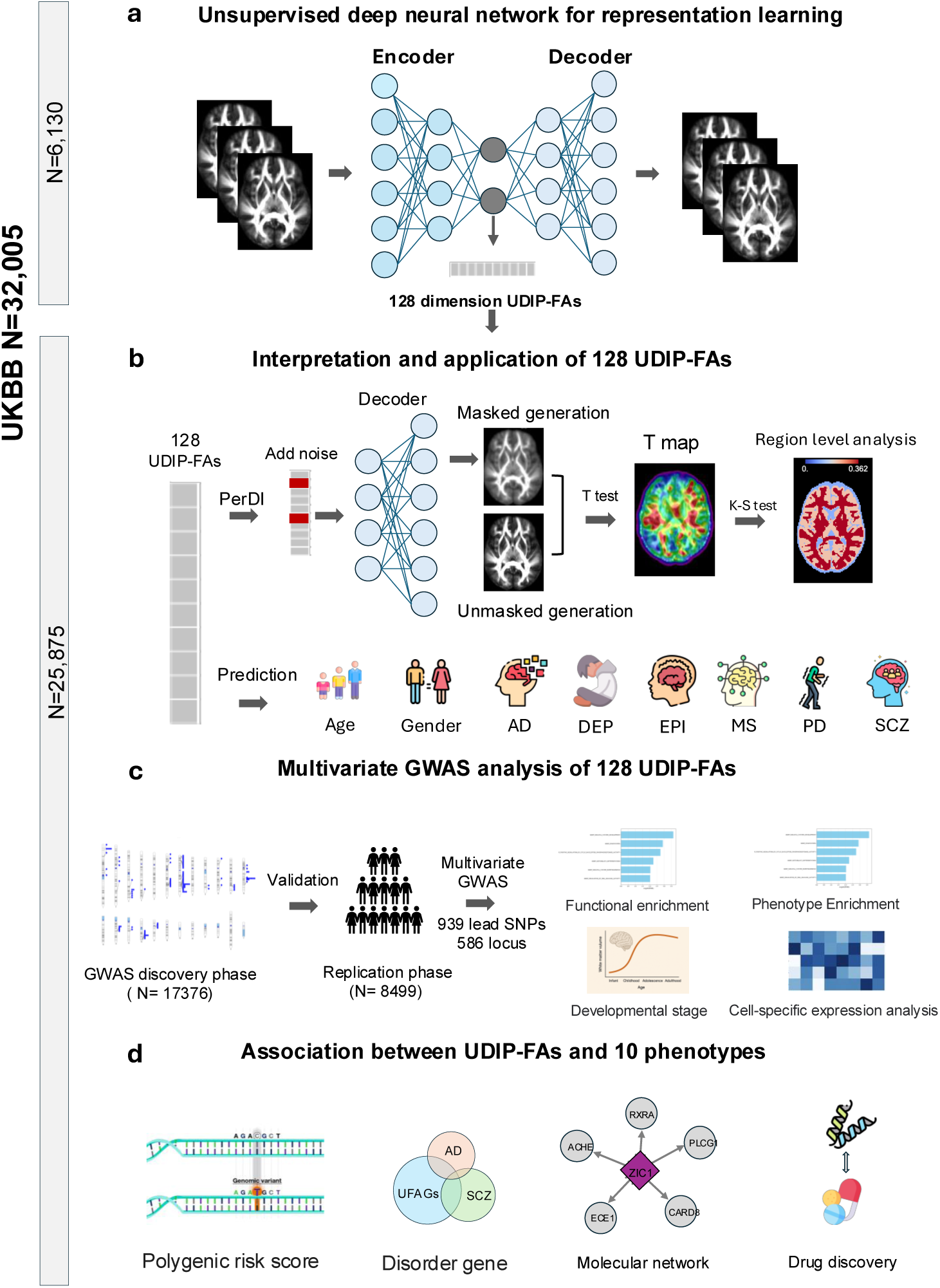
Overview of the framework for UDIP-FAs. **(a)** Model architecture for extracting UDIP-FA. **(b)** Evaluating biological meaningfulness of UDIP-FAs through interpretation and prediction of brain-related phenotypes. **(c)** GWAS discovery pipeline of UDIP-FA. **(d)** Association between UDIP-FAs and brain disorders.

### UDIP-FA characterizes the population variability of WM fiber tracts

To explore the biological relevance of the UDIP-FA features, we first assessed their ability to capture inter-individual variability in global white matter microstructure. We projected the 128-dimensional UDIP-FAs into a two-dimensional space using Uniform Manifold Approximation and Projection (UMAP) for unsupervised dimensionality reduction. Each point in the embedding was color-coded by the subject’s average whole-brain FA value. As shown in Figure 2a, UDIP-FA representations captured meaningful population-level variation, with FA values exhibiting a smooth gradient across the latent space. We next examined whether the UDIP-FAs were predictive of conventional tract-level FA measurements. Specifically, we extracted average FA values from 48 major white matter tracts defined by the UK Biobank and used multiple linear regression to assess the extent to which the 128 UDIP-FA features explained variance in each tract (Methods). Across all tracts, the UDIP-FAs significantly accounted for population-level variation (Bonferroni-corrected P < 2.2 × 10⁻¹⁶, F-test), with coefficients of determination (*R*^2^) ranging from 0.298 to 0.768 and a mean *R²* of 0.470 (Table S1).

**Figure 2:**
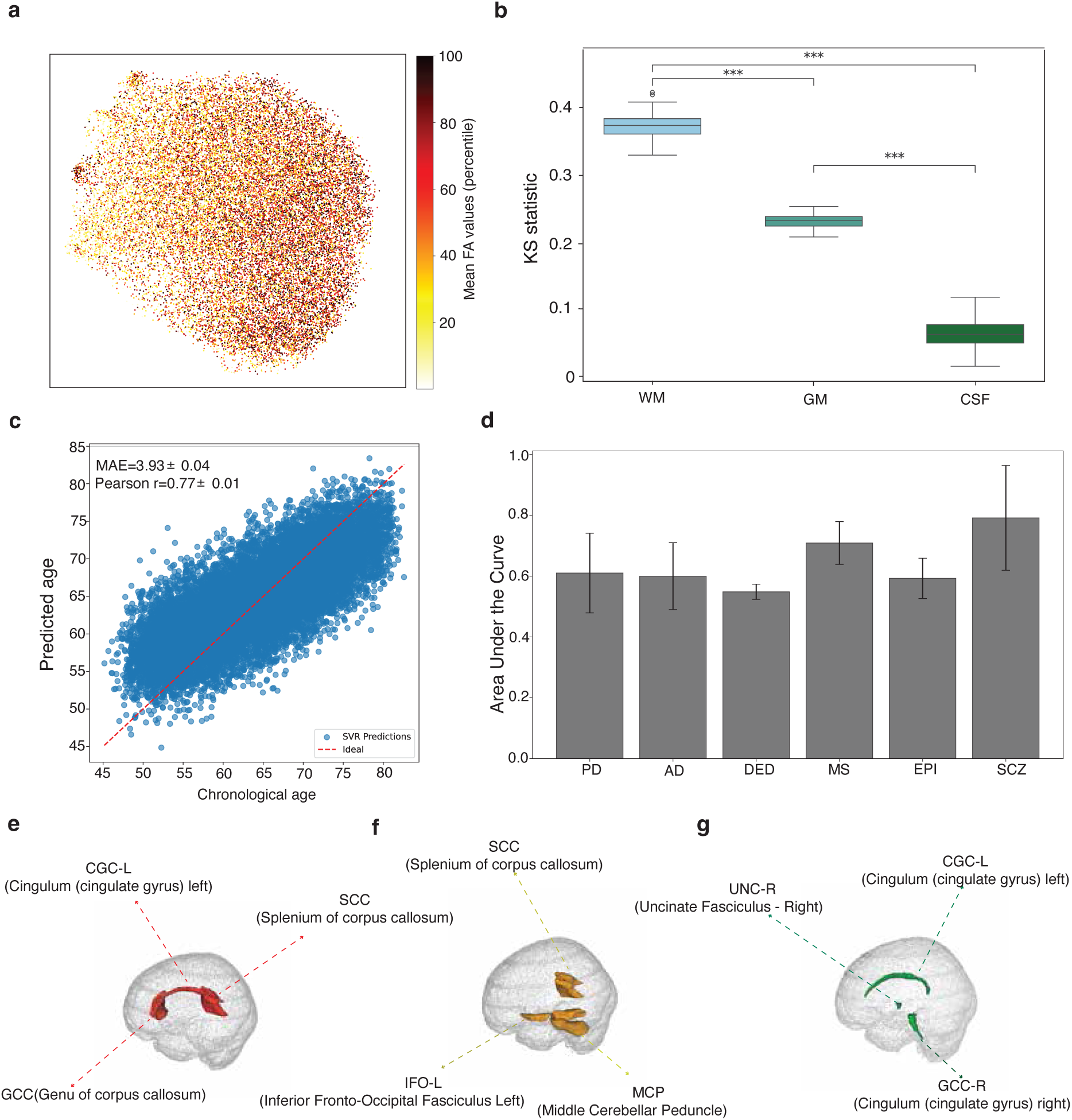
Characterization and clinical relevance of UDIPA-FA. **(a)** 2D UMAP projection of UDIP-FA representation across all subjects, colored by average mean FA value. **(b)** Kolmogorov–Smirnov (KS) statistics quantifying the distinctiveness of UDIP-FA features across three major tissue types: WM, gray matter (GM), and cerebrospinal fluid (CSF). WM region exhibited significantly higher KS scores, indicating stronger discriminative patterns (***P < 0.001, Mann-Whitney U test). **(c)** Brain age prediction using UDIP-FA features. Scatter plot shows predicted versus chronological age, with a mean absolute error (MAE) of 3.93 years and Pearson correlation of r = 0.77. **(d)** Classification performance of UDIP-FA in distinguishing individuals with various brain disorders, including Parkinson’s disease (PD), Alzheimer’s disease (AD), Depression (DEP), multiple sclerosis (MS), epilepsy (EPI), and schizophrenia (SCZ). (**e–g)** Brain regions identified as most predictive for AD (e), MS (f), and EPI (g) using feature importance of top 3 features from the GBDT classification model.

To elucidate the association between UDIPs and brain tissues, we assessed the enrichment of each UDIP’s t-map (generated via PerDI) across three primary tissue classes—WM, grey matter (GM), and cerebrospinal fluid (CSF), as well as within 81 WM fiber bundles delineated by the ICBM DTI-81 Atlas^14^ (Methods). We found that UDIPs were significantly more enriched in WM compared with GM and CSF (Figure 2b, Figure S3), and exhibited distinct enrichment patterns across different WM tracts. For example, UDIP-FA dimension 1 was notably enriched in the right uncinate fasciculus (UNC-R, mean t = 22.44, Bonferroni-corrected P = 4.99e-03, permutation test, Methods, Figure S4) and the genu of the corpus callosum (GCC, mean t = 23.40, Bonferroni-corrected P = 4.99e-03, Permutation test). Conversely, UDIP-FA dimension 2 showed significant enrichment in the right medial lemniscus (ML-R, mean t = 28.34, Bonferroni-corrected P = 4.99e-03, Figure S4) and the middle cerebellar peduncle (MCP, mean t = 27.22, Bonferroni-corrected P = 4.99e-03).

### UDIP-FA demonstrates predictive power for age, sex, and neurological disorders

To investigate the informativeness of UDIP-FA, we first evaluated its performance in sex and age prediction using 25,875 participants. For sex classification, we employed a Support Vector Machine (SVM) with five-fold cross-validation, achieving high accuracy (area under the ROC curve (AUC) = 0.987 ± 0.001). For age prediction, a support vector regression (SVR) model was used with the same training strategy. UDIP-FA demonstrated strong sensitivity to age (mean absolute error (MAE) = 3.93 ± 0.04, Pearson’s *r* = 0.77 ± 0.01; Figure 2c), outperforming a previously published study that used the traditional FA value from different WM regions^15^.

In addition, we evaluated the ability of UDIP-FA to predict the status of six brain disorders: Alzheimer’s Disease (AD), Depression (DEP), Epilepsy (EPI), Multiple Sclerosis (MS), Parkinson’s Disease (PD), and Schizophrenia (SCZ). Patients with these conditions were identified from the UK Biobank (UKB) dataset using ICD-10 diagnostic codes (field 41270, Methods). We trained a gradient-boosted decision tree (GBDT) classifier to predict diagnostic status from UDIP-FA (Methods). The results demonstrated that UDIP-FAs could be indicative across distinct brain disorders, achieving an average classification performance with an AUC of 0.642 ± 0.082 (Figure 2d). Furthermore, we assessed the feature importance of the classification model for each disorder to identify disorder-specific signatures. We found that key UDIP-FA identified by the classification model can reveal potential WM tract involvement across distinct brain disorders (Figure 2e, f, g). In AD, the critical UDIP-FA feature (*dimension_7*) was significantly enriched in the left cingulum (cingulate gyrus) (CGC-L), the genu of the corpus callosum (GCC), and the splenium of the corpus callosum (SCC), with a Bonferroni-corrected *P* < 0.05 based on permutation test results from PerDI. These WM tracts have previously been implicated in AD-related pathology^16,17^. In MS, the key UDIP-FA feature (*dimension_22*) showed significant enrichment in the left inferior fronto-occipital fasciculus (IFO-L, Bonferroni-corrected *P* = 4.99e-03,Permutation test) and the middle cerebellar peduncle (MCP, Bonferroni-corrected *P* = 4.99e-03, Permutation test), both of which have been associated with demyelination and disrupted connectivity in MS^18,19^. For EPI, the most prominent UDIP-FA feature (*dimension_116*) was significantly enriched in the right uncinate fasciculus (UNC-R, Bonferroni-corrected *P* = 4.99e-03, Permutation test). This tract has been linked to seizure propagation and altered connectivity in epilepsy^20^.

### UDIP-FA exhibited greater heritability and identified more genetic loci than conventional WM phenotypes

We used the genome-wide complex trait analysis (GCTA) software^21^ to estimate SNP-based heritability (ℎ^2^) for each UDIP-FA in an independent cohort of 25,875 individuals, quantifying the proportion of variance in each UDIP explained by common autosomal genetic variants (Methods). A total of 115 UDIP-FAs (89.8%) were significantly heritable (FDR-corrected *P* < 0.05), with heritability estimates ranging from 6.9% to 85.2% (mean = 52.0%; Figure 3A and Table S2), 39 (30.5%) UDIP-FAs showed high heritability (ℎ^2^> 0.6). The heritability of our UDIP-FAs was significantly higher than that of a previous study^12^ (mean ℎ^2^= 34.9%, P < 2.2e-16, Mann–Whitney U test, Figure 3b). This is remarkable as our UDIP-FAs are largely uncorrelated.

**Figure 3:**
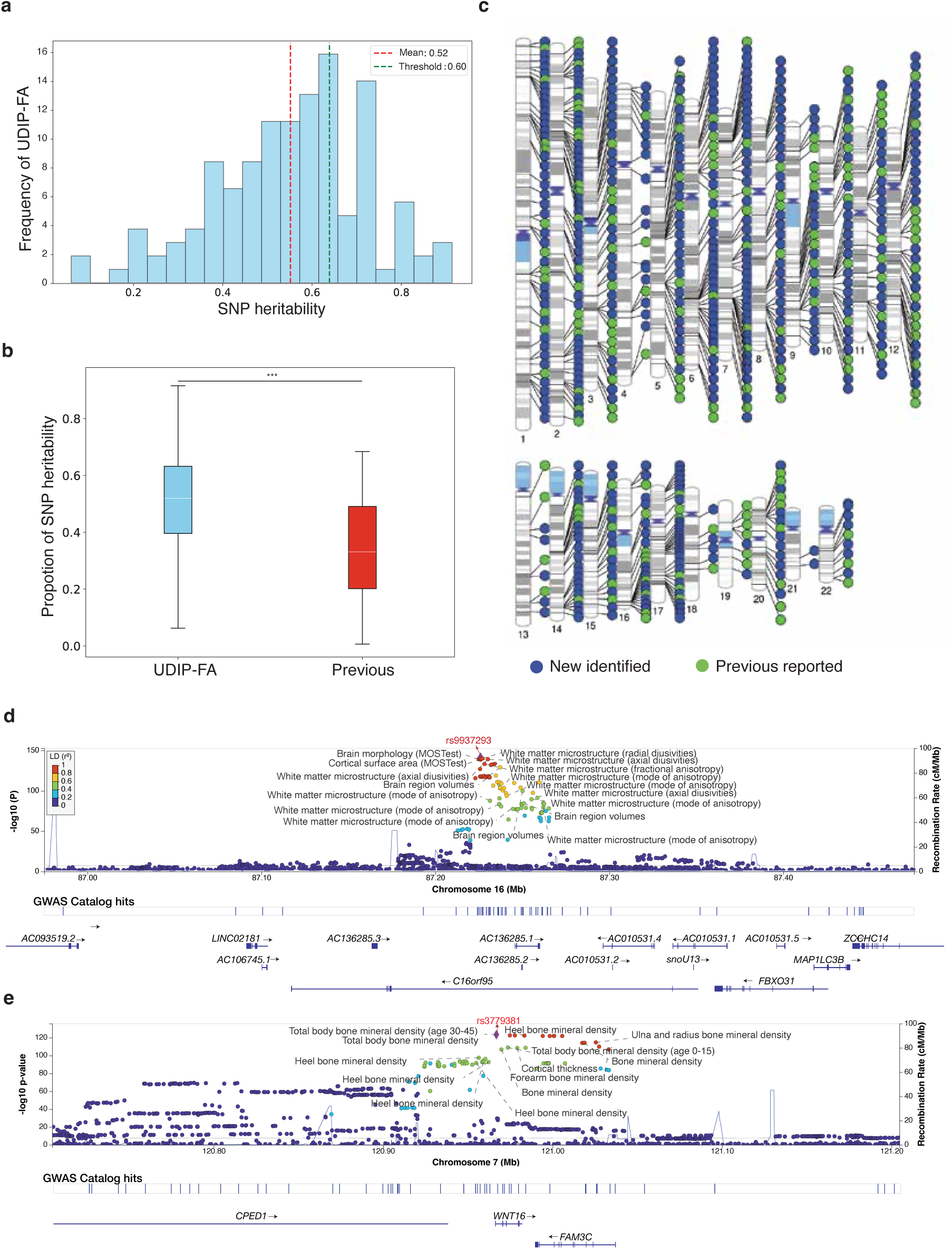
SNP heritability and the associated genomic loci of UDIP-FA. **(a)** SNP heritability of 128 UDIP-FAs. **(b)** Comparison of SNP heritability for FA between our study and the previous study. **(c)** Identified loci by meta mvGWAS. **(d)** In 16q24.1, we observed colocalization (LD R2 ≥ 0.6) between the UDIP-FA (index variant *rs9937293*) and WM microstructure-related features. (e) In 7q31.31, we observe colocalization between UDIP-FAs (index variant *rs379381*) and the total body BMD (total body BMD: index variant *rs380138*, Figure S7).

To leverage the full sample for accurately identifying variants associated with WM tracts, we employed a multi-stage GWAS analysis to ensure robust and replicable results (Methods, Figure S5). First, for the single variants, we found 128 UDIP-FA measures that showed consistent GWAS results across discovery and replication cohorts, with a high paired genetic correlation (GC = 0.969 ± 0.152; Table S3) that was evaluated by LD score regression (LDSC)^22^. Additionally, our results demonstrated significant genetic correlation with a previous GWAS of FA measures (GC = 0.294 ± 0.089)^12^, which suggesting that the UDIP-FAs reliably capture the genetic architecture of FA. Our results were able to replicate prior discoveries, with an overlap of 5 lead SNPs (4%) and 82(55.95%) genomic loci for Zhao et al.^12^, and 3 lead SNPs (1.25%) and 35 genomic loci (28.68%) for Smith et al.^23^.

Second, we applied the Joint Analysis of Multiple Phenotypes (JAGWAS) framework to perform a multivariate genome-wide association study (mvGWAS) in both the discovery and replication cohorts based on the single-variant GWAS result^24^, this approach integrated the 128 UDIP-FAs’ association signals for a SNP into a single summary statistic, enhancing the statistical power and thus the ability to detect novel SNPs associated with UDIP-FAs (Methods). Subsequently, we used FUMA to clump mvGWAS results based on linkage disequilibrium (LD) and to identify independent lead SNPs at each associated genomic locus (Methods)^25^. At a genome-wide significance threshold of *P* = 5 × 10⁻⁸, we identified 420 lead SNPs (LD *R*^2^ < 0.1) across 300 genomic loci in the discovery cohort (Figure S6a, Table S4). Among these, 182 lead SNPs (43.13%) were successfully replicated in the replication cohort (*P* < 0.05/420, Table S5, Figure S6b). In the final step, we adopted the METAL^26^ and JAGWAS to do the meta mvGWAS that combined the discovery cohort and replication cohort. By this way, at a genome-wide significance threshold of *P* = 5 ×10⁻⁸, we identified 939 lead SNPs (LD *R*^2^ < 0.1) in 586 genomic locus regions (Figure S6c, Table S6).

### Comparison with previous GWAS studies

To further investigate the biological relevance of the 939 lead SNPs identified in our meta mvGWAS, we conducted association lookups in the NHGRI-EBI GWAS Catalog^27^. The results revealed that the majority (52.29%) of lead SNPs associated with UDIP-FA had not been reported in previous GWAS (Figure 3c, Table S7). Among the 448 previously reported lead SNPs, 268 (59.82%) have been linked to brain MRI-derived phenotypes, including regional measurements of white matter microstructure such as FA and radial diffusivity (RD) in major WM tracts, as well as global WM connectome metrics^28–30^, as well as brain morphology traits (e.g., brain shape, cortical thickness, and volume) ^31,32^. Notably, SNPs in the 16q24.2 region showed strong associations with WM microstructure (Figure 3d). In addition, 25 lead SNPs overlapped with known risk variants for several neurological disorders, including schizophrenia (SCZ)^33^, Alzheimer’s disease (AD)^34^, and Parkinson’s disease (PD)^35,36^.Six SNPs were associated with cognitive functions (e.g., general cognitive ability)^37–39^,and 15 lead SNPs had previously been linked to educational attainment^38,40^. Interestingly, 42 of the identified lead SNPs were also associated with bone density traits (e.g., femoral neck and heel bone mineral density), particularly we observed shared colocalizations with femoral neck bone density and total body bone mineral density in the 7q31.31 region (Figure 3e, Figure S7). In addition, our identified SNPs also show significant association with metabolism-related traits and disease, such as High-density lipoprotein (HDL) cholesterol levels^41^ and Type 2 diabetes^42^.

### Gene-based association analysis and functional annotation

To explore the biological function associated with UDIP-FA, we first performed a gene-based association analysis using multi-marker analysis of genomic annotation (MAGMA)^43^ on the meta mvGWAS summary statistics. This analysis identified 2,201 significant gene-level associations (P < 0.05 / 19,294) between genes and UDIP-FA (Table S8), dubbed UDIP-FA associated genes (UFAGs). Among these, we replicated 126 of 413 and 56 of 137 MAGMA-identified genes from two previous studies of FA^12,28^, such as *GMNC*, *MAPT*, *VCAN*, and *PTCH*. On the other hand, we also used the proximity of physical position, eQTL, chromatin interaction to map the significant SNP to genes (Table S9, Methods), we identified 2,820 UFAGs, 1,516 (53.76%) genes were replicated in MAGMA identified UFAGs (Figure S8). In the end, we identified 3,480 UFAGs. Among them, 686 genes (19.71%) exhibit high constraint scores (pLI > 0.9), indicating intolerance to loss-of-function mutations. While the identified SNPs are not necessarily LoF variants, this enrichment suggests that many implicated genes are under strong evolutionary constraint and may play essential biological roles.

Furthermore, we examined the phenotypic associations of the UFAGs through Topgene^44^, we found that the genes associated with UDIP-FA were significantly enriched in 164 phenotypes and brain disorders. Such as WM microarchitecture (Bonferroni-corrected p < 6.27 × 10⁻⁴⁷) and brain measurements (Bonferroni-corrected P < 4.72 × 10⁻¹¹^5^, Figure 4b). Some of these genes—such as *MAPT, MPP2, LAMTOR2, RERE*, *ARHGAP27*, and *SLC41A1*—were identified as risk genes for PD (Bonferroni-corrected P < 1.988 × 10⁻⁵, Table S10).

**Figure 4:**
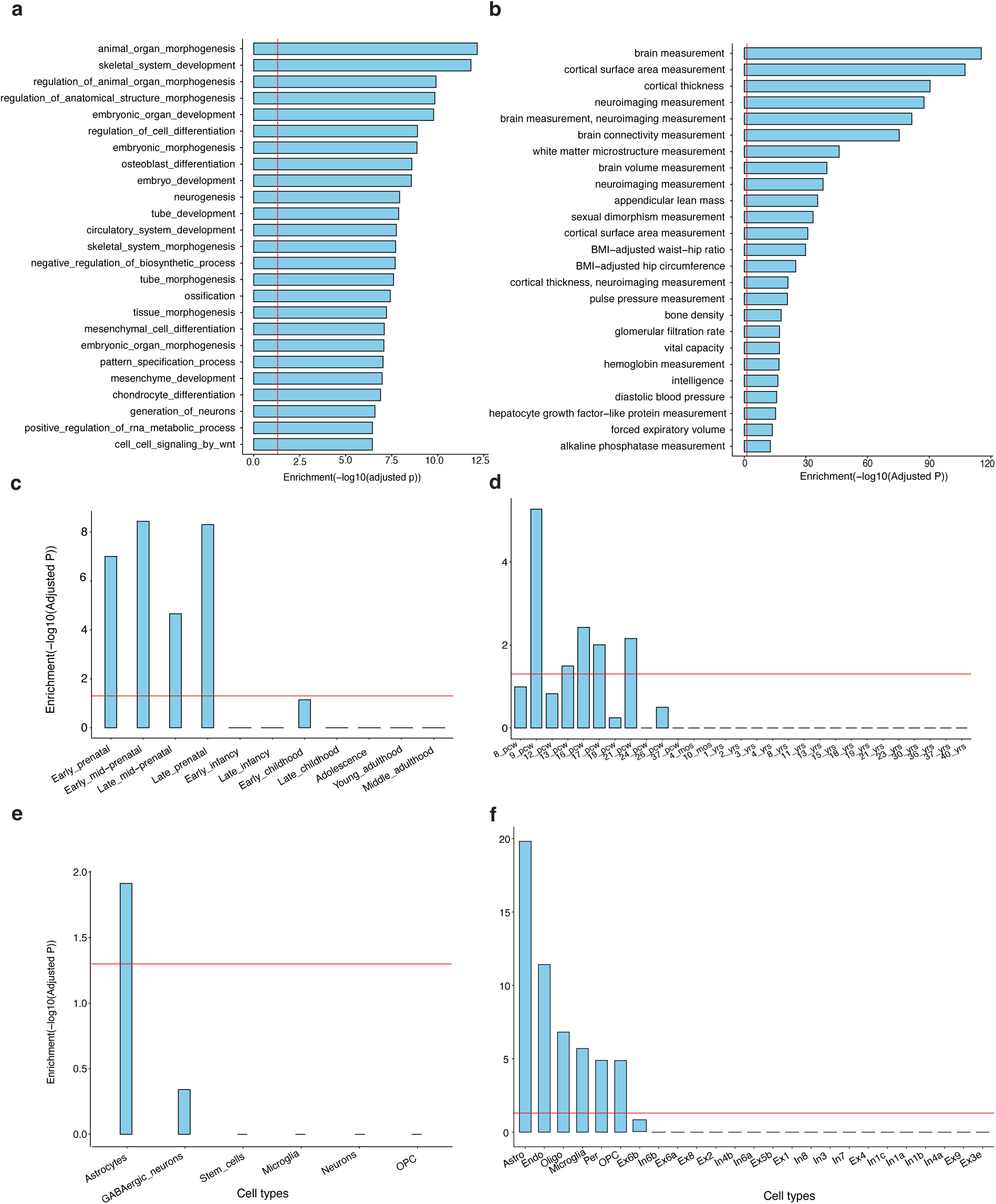
Functional annotation of UDIP-FA. **(a)** Top 25 significantly enriched Gene Ontology (GO) biological pathways. **(b)** Top 25 significantly enriched phenotypes associated with UFAGs. **(c)** and **(d)**UFAGs show prenatal up-regulation in the human brain based on BrainSpan data across 11 developmental stages or 29 age groups. **(e)** and **(f)** UFAGs are up-regulated in glial cell types based on single-cell gene expression data from the prenatal (e) and adult (f) human brain, with particularly strong expression in astrocytes. Red lines indicate the Bonferroni-corrected significance threshold of P < 0.05 within each analysis. PCW, postconceptional weeks. Astro, Astrocyte; Endo, Endothelial cell; Microglia, Microglial cell; Per, Pericyte; OPC, Oligodendrocyte precursor cell; Oligo, Oligodendrocyte; Ex6a, Ex6b, Ex6b Ex3e, Excitatory neuron subtypes from cortical layer 6, layer 5, and layer 5; Ex4, Ex2, Ex8, Ex1, Excitatory neuron from layer 4, layer 2, layer 8, and Layer 1; In1a, In1c, In1b, In4a, Inhibitory neuron subtypes from cortical layer 1 and layer 4; In3, In7, In8, Inhibitory neuron from layer 4, layer 3, layer 7 and layer 8.

In addition, we employed MAGMA to perform gene-set enrichment analysis based on p-values from the meta mvGWAS results, evaluating 17,009 predefined functional gene sets from the MSigDB database^45^. The analysis identified 196 gene sets with significant enrichment (Bonferroni-adjusted P < 0.05 for 17,009 tests; Figure 4a and Table S11). These enriched gene sets were primarily related to developmental biological processes, including “Organ Morphogenesis” (Bonferroni-adjusted P = 7.16 × 10⁻¹³) and “Neurogenesis” (Bonferroni-adjusted P = 1.17 × 10⁻⁸). Next, we conducted a MAGMA gene property analysis using human brain gene expression data from the BrainSpan database^46^, stratified by 11 life-span stages and 29 distinct age groups, to assess whether gene expression levels at specific developmental stages were associated with the strength of the correlations between genes and UDIP-FA. The results revealed that UDIP-FA-associated genes (UFAGs) were significantly enriched during the prenatal period (Figure 4c), particularly before the 21st week of gestation (Bonferroni-corrected P < 0.05; Figure 4d).

Given that WM development involves dynamic changes in the abundance and types of neuronal cells, we further examined the expression of UFAGs across different cell types at adult, using single-cell RNA sequencing data derived from the PsychENCODE ^47^. We found that UFAGs were significantly overexpressed in glial cell types during both prenatal and postnatal developmental stages (Figures 4e and 4f), especially in astrocytes, oligodendrocytes, and microglia (Bonferroni-corrected P < 0.05, Figure 4f).

### UDIP-FA demonstrates significant associations with risk of brain disorders and cognitive functions

To evaluate the association between UDIP-FA and complex diseases, we first obtained GWAS summary statistics for various diseases from published result that without sample overlap with the UK Biobank dataset used in our study (Methods). We then computed the polygenic risk scores for each disease using PRS-CS ^48^.To assess the associations between the 128 UDIP-FA features and polygenic risk scores (PRS) for 11 traits(Table S12), we conducted canonical correlation analysis (CCA), adjusting for relevant covariates (Methods). Our results revealed significant associations between UDIP-FAs and several brain disorders (Figure 5a), including multiple sclerosis (MS; canonical correlation coefficient (Rc)= 0.111, Bonferroni-corrected P = 2.95× 10^-5^, F test), schizophrenia (SCZ; Rc = 0.110, Bonferroni-corrected P = 6.06×10^-5^, F test), and amyotrophic lateral sclerosis (ALS; Rc = 0.104, Bonferroni-corrected P = 3.77× 10^-3^, F test). Notably, both MS and ALS are neurodegenerative disorders with well-established white matter involvement, suggesting that UDIP-FA may capture disease-specific microstructural alterations in white matter with a certain degree of biological specificity.

**Figure 5:**
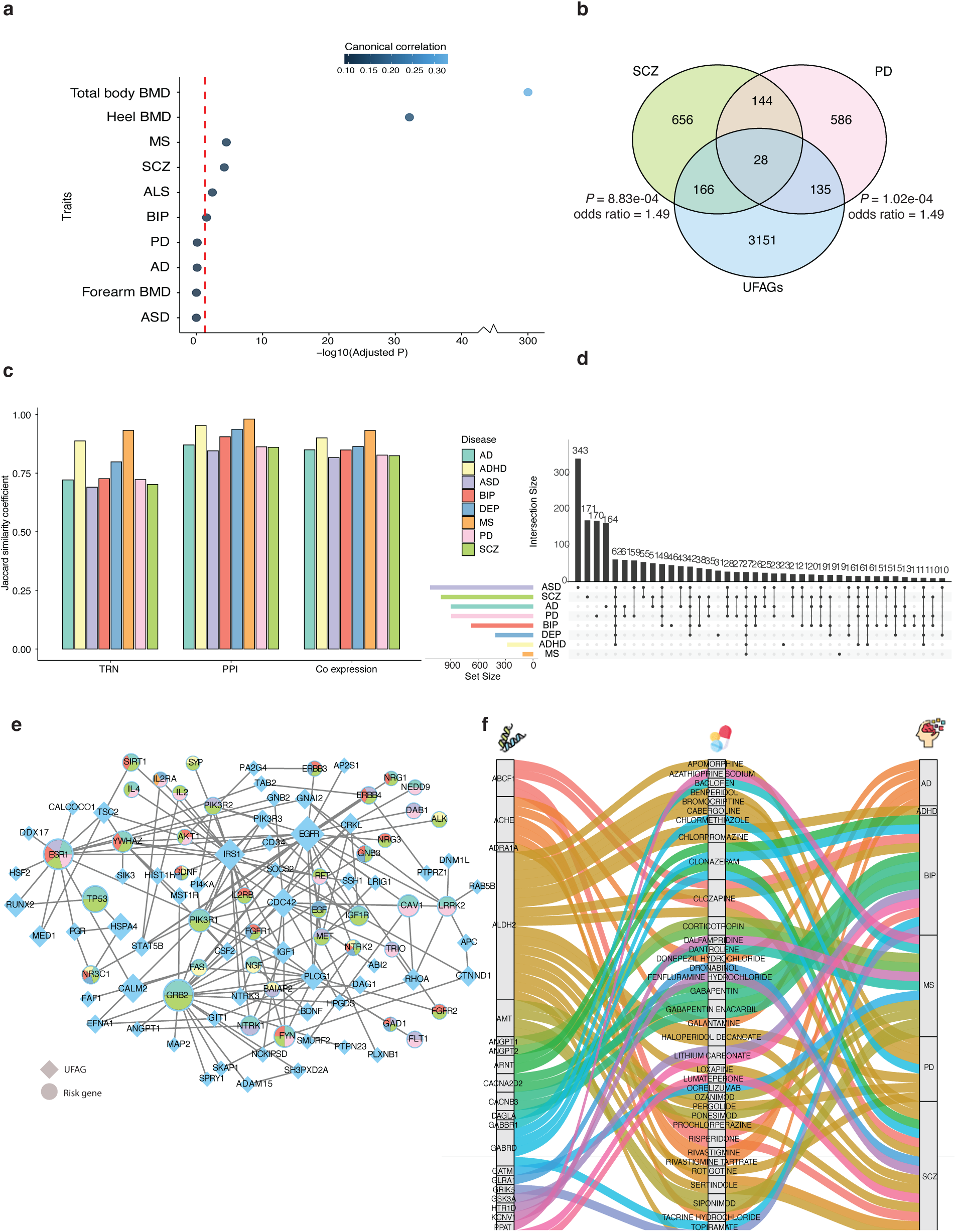
Associations between UDIP-FA and various diseases at both phenotypic and molecular levels. **(a)** Bubble plot showing significant associations between UDIP-FA and the polygenic risk scores (PRS) of brain disorders, as well as bone mineral density (BMD). Red lines indicate the Bonferroni-corrected significance threshold (*P* < 0.05) within each analysis. **(b)** Venn diagram illustrating significant enrichment of UFAGs within the risk gene sets of schizophrenia (SCZ) and Parkinson’s disease (PD). *P*-values are Bonferroni-corrected. **(c)** Enrichment of UFAGs with risk genes for various brain disorders within molecular networks. Enrichment is quantified using the Jaccard distance index, where higher values indicate greater similarity. Analyses are based on transcriptional regulatory networks (TRNs), protein–protein interaction (PPI) networks, and gene co-expression networks derived from the human brain (Methods). **(d)** Overlap of protein interacting with UFAGs in brain-active PPI networks for multiple brain disorders. **(e)** Network visualization of pivotal hub modules for UFAGs in the PPI network. Diamonds represent UFAGs, circles denote disease risk genes, and pie chart colors indicate gene overlap across different brain disorders. **(f)** Tripartite network connecting subset of UFAGs, drug compounds, and brain disorders. This alluvial plot depicts associations among UFAGs (left), drug compounds (middle), and major neuropsychiatric and neurodegenerative disorders (right). Colored flows represent known gene–drug–disease relationships curated from established databases (Methods). AD, Alzheimer’s disease; ADHD, Attention Deficit Hyperactivity Disorder; BIP, Bipolar Disorder; DEP, Depression; MS, Multiple Sclerosis; PD, Parkinson’s Disease; SCZ, Schizophrenia; ASD, Autism Spectrum Disorders.

Additionally, UDIP-FAs were strongly associated with bone density–related phenotypes, such as total body bone mineral density (BMD; Rc = 0.332, Bonferroni-corrected P = 2.12× 10^-307^, F test) and heel BMD (Rc = 0.156, Bonferroni-corrected P = 7.38×10^-33^, F test).

### Genetic pleiotropy of UFAGs and risk genes in various brain disorders

Since UDIP-FA shows significant associations with various brain diseases and demonstrates potential for disease classification, we sought to investigate the underlying molecular mechanisms between WM variations captured by UDIP-FA and brain disorders. To this end, we collected eight risk gene sets associated with different brain disorders (Methods, Table S13) and examined their overlap with UFAGs. Our analysis revealed that UFAGs intersect with multiple disease-associated gene sets and show particularly significant enrichment for risk genes of SCZ (Bonferroni-corrected P = 8.83 × 10⁻⁴, Fisher’s exact test) and PD (Bonferroni-corrected P = 1.02 × 10⁻⁴, Fisher’s exact test) (Figure 5b).

To further explore potential biological relationships between UFAGs and disease-associated genes, we analyzed interactions between UFAGs and the eight risk gene sets across three brain-specific molecular networks (Methods), we found UFAGs exhibit stronger enrichment with risk genes in protein-protein interaction (PPI, Jaccard distance (*J*) = 0.90) and co-expression networks (*J* =0.85) compared to transcriptional regulatory networks (TRNs, *J* = 0.77, Figure 5c). This likely reflects their primary roles of UFAGs as structural or effector genes—such as those involved in myelination, axonal guidance, or cytoskeletal organization—rather than upstream transcriptional regulators. Several UFAGs, including *MOG, MOBP, SOX10,* and *RTN4*, are well-established contributors to oligodendrocyte development, axon guidance, and myelination. These genes demonstrate strong connectivity in protein-protein interaction and co-expression networks, underscoring their roles in WM biology, while being less prominent in transcriptional regulatory hierarchies^46^.

To investigate the impact of UFAGs on various brain disorders, we first assessed the pleiotropy of disorder-associated risk genes modulated by UFAGs within the brain-activated protein–protein interaction (PPI) network^49^(Table S14). Our analysis revealed that 55.82% of these risk genes interacting with UFAGs were implicated in at least two brain disorders (Figure 5d). Intriguingly, we identified *EGFR* as a major hub module (ranked by degree, Figure 5e), interacting with numerous neurodegenerative disease risk genes. Proteins associated with *EGFR* are significantly enriched in RTK signaling pathways, which have been directly implicated in the regulation of fractional anisotropy (FA): RTK signaling promotes oligodendrocyte precursor cell (OPC) proliferation and differentiation, and its disruption impairs myelination and reduces FA, as demonstrated in both genetic and pharmacological models ^50,51^.

We further examined the interactions between UFAGs and disorder-related risk genes within the human brain’s co-expression and TRN networks. Consistent with the brain PPI network findings, 76.81% and 48.22% of the risk genes interacting with UFAGs were associated with at least two brain disorders in the co-expression and TRN networks, respectively (Figures S9 and S10).

To identify potential therapeutic targets, we queried the Drug–Gene Interaction Database (DGIdb)^52^ to extract stable associations between UFAGs and approved drugs (Methods), focusing specifically on brain-related disorders. This analysis revealed that 44 UFAGs interact with 87 drugs used to treat 55 distinct brain-related diseases (Table S15). We further examined the connections between UFAGs and the eight brain disorders incorporated in our drug–gene Interaction network analysis (Figure 5f) and found that many UFAGs serve as known drug targets. For example, *ACHE* is a target of anticholinesterase inhibitors such as galantamine and rivastigmine, which are well-established treatments for Alzheimer’s disease. Similarly, *ALDH2* is targeted by antipsychotic drugs, including clozapine and haloperidol decanoate, both of which are used in the treatment of SCZ and PD. These findings highlight a subset of potentially druggable UFAGs and underscore shared pharmacogenomic targets across multiple brain disorders.

### Comparison of UDIPs across different brain imaging modalities

Brain images acquired through different modalities provide complementary insights^53^; however, the interrelationships among these modalities remain inadequately characterized. To investigate these relationships, we conducted a comprehensive extraction and analysis of whole-brain images from three modalities—UDP-FA, UDP-T1, and UDP-T2—using data from the UKB. Cross-modality relationships were evaluated using CCA applied to 128-dimensional UDIP representations (Figure 6, Methods). Results showed that FA (0.34 ± 0.06) were more heritable than T1(0.26 ± 0.03) and T2(0.25 ± 0.04) that were evaluated by LDSC^13^. T1 and T2 shared the highest mutual explainability (CCA explained variance: 91%), whereas T1 was better explained by FA (74%) than T2 (69%). Building upon our prior investigation into the genetic architecture of UDP-T1 and UDP-T2, we extended the analysis to genetic correlation across all three modalities (Methods). The genetic correlation between T1 and T2 was stronger than either’s relationship with FA. Furthermore, loci identified across different models revealed a consistent pattern: T1 and FA shared 374 loci (80.09% overlap), while T2 shared 399 loci (86.18%) with the same set.

**Figure 6:**
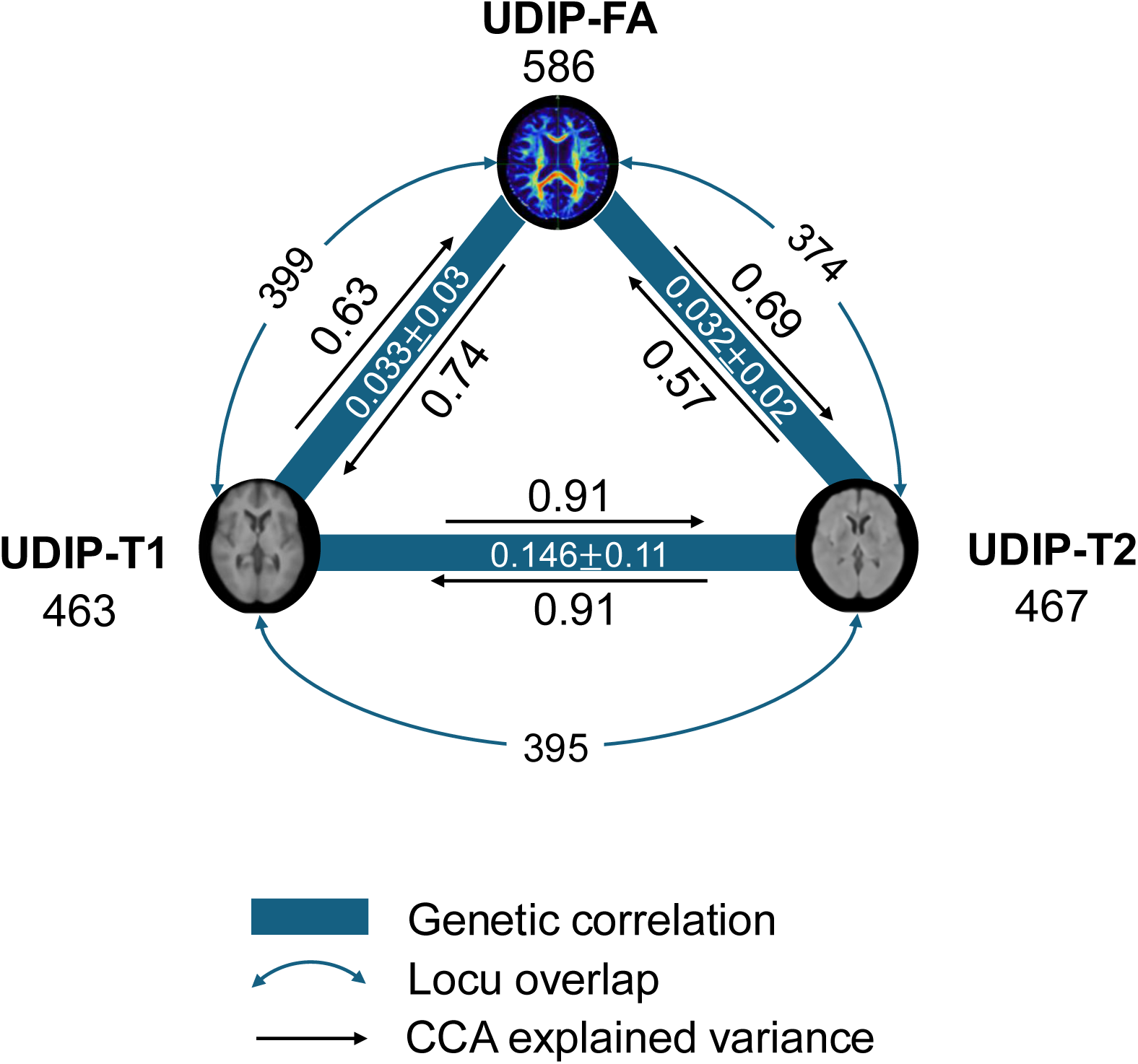
Cross-modality relationships among diffusion, T1-weight, and T2-weight MRI. The number below each modality label denotes the number of genome-wide significant loci identified in the multivariate GWAS. Blue solid lines represent genetic correlations estimated between each modality pair; the corresponding values are indicated alongside the lines. Black curved arrows denote canonical correlation analysis (CCA)–based explained variance between modality-specific UDIP representations. Dashed gray arrows indicate the number and proportion of overlapping loci between modality pairs.

## Discussion

In this study, we developed UDIP-FA, an unsupervised deep representation of WM microstructure and demonstrated its effectiveness in capturing biologically and genetically meaningful brain variation. UDIP-FA outperforms traditional FA phenotypes in representing population-level WM architecture, with strong predictive power for age, sex, and multiple brain disorders. UDIP-FA traits show high heritability and GWAS of UDIP-FA identified many novel and replicable loci. The associated genes (UFAGs) are enriched for neurodevelopmental and show significant overexpression in glial cells. We further found that UFAGs are pleiotropic, overlap significantly with brain disorder risk genes, and interact with known drug targets, supporting their translational potential. UDIP-FA offers a robust and interpretable framework to advance our understanding of WM biology and its role in brain health and disease.

Compared with conventional FA phenotypes derived by hand-crafted ROI or tract-based approaches, UDIP-FA extracted via an unsupervised 3D convolutional autoencoder, which better encodes inter-regional WM relationships, as it directly uses FA maps as input, and passes them through a minimal encoder–decoder bottleneck. This design ensures maximal information compression while preserving WM-relevant patterns. We demonstrate that UDIP-FA supports higher-accuracy brain age prediction using only simple machine learning models than before^15^. Furthermore, although our network was never trained on disease labels, UDIP-FA naturally highlights disease-critical WM tracts, such as those implicated in AD (e.g., cingulum, corpus callosum), and shows significant correlations with cognitive scores and genetic risk factors for multiple brain disorders. This finding aligns with studies showing that unsupervised learned representations can uncover subtle pathology-driven patterns and correlate with established AD biomarkers^54^. Our findings indicate that UDIP-FA serves as a robust and unbiased biomarker, capturing rich microstructural and network-level features crucial for assessing WM integrity across ageing and disease. These representations can augment conventional image-derived phenotypes in GWAS and clinical prediction, ultimately improving both interpretability and biomarker sensitivity in neurodegenerative and WM disorder research. Additionally, this study demonstrates that UDIP-FA captures significant associations in disorders such as MS and ALS, where WM abnormalities are known to play a key pathophysiological role. These findings imply that UDIP-FA is sensitive not only to anatomical distinctions but also to functionally driven structural alterations in the brain.

Multi-stage GWAS of UDIP-FA have identified many novel loci that reveal the genes relevant to the biological process of myelination and glial cell function associated with WM microstructure. The locus tagged by *rs4239889* is located near *SOX10*, a well-known master regulator of oligodendrocyte differentiation that drives the expression of myelin-related genes and ensures oligodendrocyte survival^55,56^. *rs1768234* maps near *MOBP*. *MOBP* encodes a structural component of compact CNS myelin, marking mature oligodendrocyte functionality, and it plays a crucial role in myelination and axonal integrity that influencing WM microstructure. Other loci include *rs34580448* (near *VCAN*), and *rs11259505* (regulated the *ADAMTSL4* by eQTL). *VCAN,* and *ADAMTSL4* are extracellular matrix-related genes that regulate pericellular and microfibrillar scaffolding, respectively, contributing to axonal stability and myelination processes that underlie WM microstructural integrity^10,57^. Additionally, some UIDP-FA related loci that are members of the WNT gene family, such as *WNT3A*, *WNT5A* acting through canonical Wnt/β-catenin signaling, are essential regulators of WM myelination, with dysregulated pathway activity in oligodendrocyte precursor cells shown to delay developmental myelination and remyelination in the CNS^58^. Additionally, *WNT16*, along with *WNT4* and *WNT5A*, are strongly implicated in bone mineral density (BMD) and skeletal strength, supported by GWAS and functional studies linking *WNT16* to cortical thickness, fracture risk, and osteoblast differentiation^59^. These findings suggest that Wnt signaling may represent a shared molecular mechanism underlying both WM integrity and BMD, potentially linking neurodevelopmental and skeletal pathways through common genetic regulation.

UFAGs characterize the potential underlying molecular regulation of WM in different brain diseases and have the potential to be a drug target. In the brain TRN, our findings suggest that the transcription factor *MEF2C* plays a central role in the shared genetic architecture between WM integrity and major psychiatric disorders^60^. Specifically, we show that *MEF2C* regulates multiple pleiotropic risk genes implicated in both BIP and SCZ, such as *GRIN2A*, *MCHR2*, and *YWAH*. Loss-of-function variants in *MEF2C* itself are known to cause developmental delay, intellectual disability, and autism, and are typically associated with neuroimaging findings such as hypoplastic corpus callosum, mild thinning of the cortical white matter, delayed myelination, and mild undermyelination^61,62^. These genes are involved in synaptic signaling, neurotransmitter regulation, and intracellular signaling pathways that are essential for neurodevelopment and cognitive function. For example, *GRIN2A* encodes an NMDA receptor subunit critical for synaptic plasticity and has been linked to SCZ and cognitive dysfunction^63^. Given that *MEF2C* functions as a transcription factor involved in the regulation of synaptic and neurodevelopmental genes^64^, it is plausible that its regulatory influence extends to pleiotropic risk genes such as *GRIN2A*, *MCHR2*, and *YWAH*, which are implicated in BIP and SCZ. In addition, the UFAG, *ANGPT1*, serves as a convergent downstream target of multiple risk transcription factors implicated in distinct brain disorders. Notably, in the context of MS, *ANGPT1* is regulated by key immune-modulatory TFs such as *PPARG* and *STAT3*, where *PPARG* mediates anti-inflammatory responses and ameliorates disease progression, while *STAT3* promotes pro-inflammatory Th17 differentiation and neuroinflammation^65^. Functionally, *ANGPT1* plays a critical role in maintaining blood-brain barrier (BBB) integrity and vascular stabilization, and its dysregulation has been implicated in BBB breakdown and increased neurovascular permeability observed in MS lesions^66^, We also found that it interacts with the MS treatment drug corticotropin, which suggests that *ANGPT1* may serve as a potential therapeutic mediator or downstream effector in modulating vascular responses during immunomodulatory treatment in MS.

Our findings delineate a hierarchy of modality integration across brain imaging and genetics: T1 and T2 modalities form a tightly integrated pair, both phenotypically and genetically (average cross-modality correlation and overlapping UDIP-derived loci)^13^, while FA captures complementary microstructural information (e.g., voxel-wise aging effects distinct from T2 contrasts) ^67^. This layered understanding has several implications. First, it recommends selecting structurally similar modalities (e.g., T1–T2) when designing imaging-genetics studies to maximize shared signal, as deep-embedding GWAS showed enhanced overlap and statistical power in these modalities. Second, discovering modality-specific loci enriches our ability to link particular genes with distinct neurobiological substrates— FA-associated loci highlight genes involved in myelination or fiber tract integrity^68^, whereas T1/T2-associated loci are more likely to implicate genes involved in tissue composition, cellular organization, or iron homeostasis, reflecting their sensitivity to cell density and regional iron concentration^69,70^, as reflected by loci uniquely identified from UDIP features. Lastly, these insights can refine trait mapping in neurodegenerative and developmental disorders by targeting imaging features with maximal genetic sensitivity, leveraging modality-specific genetic signals uncovered in multivariate GWAS frameworks.

We demonstrate that UDIP-FAs show strong correlations with traditional FA phenotypes, enhancing their interpretability. These patterns extend beyond predefined, single-region volumetric features such as IDPs, which presents challenges for interpretability. Although our PerDI approach offers a partial solution, further improvements are needed. Additionally, our UDIP-FA GWAS was conducted exclusively on European populations, limiting its ethnographic generalizability. Future studies will aim to validate the robustness of these findings across diverse populations.

In summary, our study is the first to apply unsupervised deep representation learning to FA maps that derived from DTI, leading to the development of UDIP-FA. We demonstrate that UDIP-FA captures a more informative and functionally relevant representation of WM microstructure, enabling deeper insights into its genetic architecture and disease associations. Importantly, UDIP-FA reveals previously unrecognized links between WM integrity and brain disorders, and highlights potential therapeutic targets, suggesting its promise as a bridge between imaging biomarkers and pharmacological intervention.

## Methods

### FA maps acquisition and preprocessing

UK Biobank (UKB) was chosen for this study because it represents the largest publicly available brain imaging dataset, uniformly processed through standardized pipelines, ensuring consistency and comparability across subjects^71^. We used data from 30789 UKB individuals of British ancestry (self-reported ethnic background, data-field 21000) that have the diffusion MRI image data. The diffusion MRI data in UKBB were primarily acquired using Siemens Skyra 3T MRI scanners operating VD13A SP4, equipped with a standard Siemens 32-channel RF head coil. The diffusion-weighted images (DWIs) utilized a multi-shell diffusion sequence, optimized for consistent quality across multiple imaging centers^67^. To maximize generalizability and reduce feature engineering, we followed the straightforward preprocessing pipeline provided by the UKBB imaging team, primarily utilizing the FMRIB Software Library (FSL; https://www.fmrib.ox.ac.uk/ukbiobank/). Key preprocessing steps provided by UKBB include correction for motion and eddy current distortions using FSL’s eddy tool, diffusion tensor fitting using FSL’s DTIFIT, and generation of bias-field-corrected FA maps^72^. Subsequently, all FA maps were spatially normalized to the MNI152 space with the affine part of the transformation provided by UKBB using FSL FLIRT. Normalization ensured the standardization of head sizes and alignment of brain structures across subjects, while preserving relevant structural deformation information. All individuals were aged between 40 and 80 years and the proportion of females was 52.7%.

### Unsupervised Deep Neural Network for Representing FA maps

For obtaining the UDIP-FA, we adopted our previously proposed model - Deep 3D convolutional autoencoder to obtain the 128-dimensional phenotype. A separate model was trained for FA maps. The architecture was implemented using PyTorch and trained with the PyTorch Lightning framework. To obtain representations of the whole brain, we take the FA maps linearly registered to the MNI-152 atlas as input. The model consisted of 5 convolutional encoder blocks, a linear latent space of 128 dimensions, 5 convolutional decoder blocks. It has 138.12 million parameters (Figure S1)^13^. The output reconstructed image is of the same size as the input MRI (182 × 218 × 182).

Our model employs a standard convolutional autoencoder without skip connections to preserve maximal information through the bottleneck, enabling each latent vector (UDIP) to capture global brain morphology. Unlike prior approaches that reduce resolution, extract patches, or focus only on WM tracts, we process full-resolution, whole-brain MRIs, maintaining anatomical completeness. Leveraging the large UK Biobank dataset, our unsupervised framework is the first of its kind to derive comprehensive, scalable imaging phenotypes (UDIP-FA) for brain-wide GWAS.

For training this model, a dataset of 6130 images from subjects of mixed ethnicities was chosen as the model development set. We randomly split the dataset into training (75%) and validation (25%) sets, using the latter to tune hyperparameters and select the best-performing model checkpoint based on validation loss. Our autoencoder uses a voxel-wise regression framework with no output activation, optimized via masked mean squared error focused solely on brain regions. Models were trained for 75 epochs using the Adam optimizer and a well-tuned learning rate of 1.4 × 10^-4^, with training accelerated by two NVIDIA A100 GPUs. This setup enabled efficient and anatomically precise image reconstruction for FA maps.

### Interpretation and application of UDIP-FA

To interpret spatial representations of each of the 128 UDIP-FAs, we developed PerDI—a perturbation-based approach. For a given UDIP-FA, Gaussian noise (*σ*) is added while other dimensions remain unchanged. The original decoder reconstructs images from the perturbed and unperturbed representations for 500 randomly selected individuals. A paired t-test between these two sets of images yields an absolute t-map highlighting brain regions associated with the target UDIP. The resulting map is smoothed with a Gaussian filter (*σ* = 3) to mitigate registration imperfections.

To explore the regional enrichment of UDIP-FA, we used the brain tissue segmentation atlas^73^ and ICBM DTI-81 atlas^14^ to annotate t-maps. Voxels were ranked by t-values in descending order. For each atlas-defined region, a normalized Kolmogorov–Smirnov (K–S) statistic was computed to quantify regional enrichment. The K–S curve is defined as 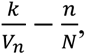 where *k* is the number of voxels from a region among the top n ranked voxels, *V_n_* is the number of voxels in the region of atlas, and *N* is the total number of voxels. The K-S value is the maximum value along the K-S curve. A higher K–S value indicates stronger representation of the region by the corresponding UDIP-FA. On the other hand, to evaluate the statistical significance of voxel-wise effects within the atlas’ predefined regions-of-interest (ROIs)s for UDP-FA, we employed a non-parametric permutation test. Specifically, the test was conducted using an ROI mask *M* partitioning the WM into *R* discrete anatomical regions, each denoted as *Vr* = {*v*: *M*(*v*) = *r*}, where *r* ∈ {1, …, *R*}. First, for each ROI *r*, the observed statistic 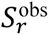 was calculated as the median of the absolute voxel-wise t-values derived from the statistical parametric map (T-map) of PerDI: 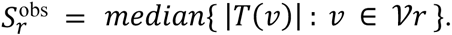 Subsequently, a permutation-based null distribution was constructed by randomly shuffling the observed t-values within the combined ROI mask, thereby breaking any true spatial associations. For each permutation i of *n*perm = 5000, a permuted statistic 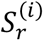 was computed for each ROI. Raw p-values were then estimated as, 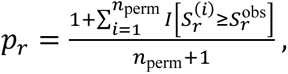 where *I*(⋅) denotes the indicator function. To correct for multiple comparisons across ROIs, bonferroni correction was applied. These spatial enrichments were then used to associate UDIP-specific brain regions with brain disorders.

### UDIP for predicting disease status and associated with cognitive function

To get the disease status of the UKB samples, we used the ICD-10 code (field: 41270) to obtain the patients for different brain disorders, including AD (F00, G30), PD (G20), SCZ(F20), DEP(F32), and MS(G35). And obtain the cognitive function of the fluid intelligence score (field: 20016), prospective memory (field: 20018), and mean time to correctly identify matches (field: 20023).

To evaluate the classification performance of UDIP-FA features across various brain disorders under imbalanced sample conditions (e.g., 26,000 controls vs. 216 MS cases), we adopted a five-fold stratified cross-validation framework combined with stochastic down-sampling of the majority class. Specifically, for each cross-validation fold, the dataset was split into training and testing sets while preserving the original class distribution using stratified sampling.

Within each training set, we randomly down sampled the majority class (controls) to match the number of minority class samples (e.g., MS cases), ensuring class balance during model training. Formally, given two classes A and B, where |A| > |B|, a random subset *A*’ ⊂ *A* was sampled such that |A’| = |B|, and the balanced training data consisted of *A*’ ∪ *B*. The held-out testing fold was kept at its original class distribution to fairly assess model generalization.

For the classification task, we employed gradient boosted decision trees (GBDT) implemented using the LightGBM Python package with default hyperparameters^74^. Model performance was evaluated on each test fold using the area under the receiver operating characteristic curve (AUC), and the mean AUC across the five folds was reported. Feature importance was determined using LightGBM’s gain-based ranking, averaged across folds to identify robust predictive markers.

### Genetic data preprocessing and association analysis

Genetic analyses were performed on individuals from the UKB with European ancestry, who also had genotypes and diffusion MRI image data. Standard quality control procedures were then applied to the UKB v3 imputed genetic data^75^. These procedures included the following steps: (1) exclusion of individuals with failed genotyping, abnormal heterozygosity status, or withdrawn consents; (2) removal of participants genetically related—up to the third degree—to another participant, as inferred by kinship coefficients implemented in PLINK^76^; (3) elimination of variants with a minor allele frequency below 0.01%; (4) removal of variants with a genotype missing rate exceeding 10%; (5) exclusion of variants failing the Hardy-Weinberg equilibrium test at the 1e-07 level; 6) elimination of variants with an imputation INFO score below 0.8. Post quality control, we retained 30,789 individuals and 8,931,083 variants.

For genome-wide association analysis (GWAS) of the UDIP-FA, we adopt the multiple-stage GWAS analysis for the UDIP-FA (Figure S6). 1) We first divided the samples into discovery and replication cohorts, followed by conducting a genome-wide association study (GWAS) on the 128-dimensional UDIP traits separately for each cohort using mixed linear models implemented in GCTA^21^. Covariates included age (field ID 21003), age squared, sex (field ID 31), interaction terms (sex × age, sex × age²), the first 10 genetic principal components (field ID 22009), head size (field ID 25000), head position in the scanner (field IDs 25756–25758), scanner table position (field ID 25759), assessment center location (field ID 54), and date of assessment (field ID 53).To evaluate the stability of the GWAS results, we conducted LDSC^77^ for the whole GWAS, which yielded intercept values close to 1, indicating that the observed inflation in test statistics is likely due to polygenicity rather than confounding factors. We also validated the reliability of our findings by comparing results between the discovery and replication cohorts. 2). To assess whether single genetic variants influence multiple UDIP dimensions, we employed JAGWAS^24^ to perform multivariate GWAS analyses in both the discovery and replication cohorts. 3) We used the method METAL^26^ to perform the meta-analysis to integrate discovery and replication cohort of 128 GWAS of UDIP-FA to improve the detection effectiveness of our SNPs. 4) Finally, we applied the JAGWAS^24^ to perform the multivariate GWAS analysis on 128 meta GWAS summary statistic, this method to integrate the 128 meta GWAS of UDIP into a single multivariate GWAS summary statistic. The *P*-value threshold for selecting the significant multivariate GWAS variants is 5e-08.

### Prioritizing UFAGs and gene expression analysis

In the study of the UDIP-FA meta mvGWAS, MAGMA (version 1.08)^43^ was used to perform a gene-based association analysis for 19,218 protein-coding genes. The default MAGMA parameter settings were applied, with a zero-window size around each gene. Subsequently, FUMA functional annotation and mapping analysis were performed, which involved annotating variants with their biological functionality and linking them to candidate target genes through a combination of eQTL and 3D chromatin interaction mappings. Brain-related tissues/cells were selected for all options, and default parameters were used.

In addition, to explore the tissue/cell specific and developmental stage using the MAGMA based on tissue expression data and cell expression data. We used MAGMA to test whether cell or tissue-specific gene expression levels predict the strength of GWAS associations for UDIP-FAs. This gene-property analysis assesses continuous gene characteristics (e.g., expression in a specific tissue) rather than predefined gene sets. After performing gene-level association analysis, each gene’s p-value (*p_g_*) was converted to a Z-score:

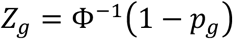

where Φ^-1^ is the probit (inverse normal) function. We then modeled *Z_g_* via linear regression:

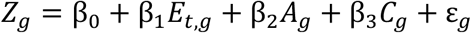

Where *E_t,g_* is expression level of gene g in tissue t, *A_g_* was average expression across all tissues (controls for baseline), *C_g_* is covariates (e.g., gene size, SNP density); *β*_1_was the coefficient testing whether tissue t expression is positively associated with gene-level association. A significantly positive *β*_1_ indicates that higher expression in tissue t is associated with stronger genetic effects.

For tissue and cell type expression data, we used BrainSpan^46^ and PsychENCODE ^47^. Then we applied Bonferroni correction across all tested tissue or cell types.

### Computing polygenic scores and correlation with UDIP-FA

The polygenic score (PGS) represents an estimate of an individual’s genetic risk for a given trait. To estimate the association between UDIP-FA and genetic risk for different traits, the polygenic score for complex traits was generated using PRS-CS^48^ by default parameters, which based on genetic data of Europeans from UKB with the collected GWAS data for 11 complex traits (Table S12), including SCZ, BIP, autism spectrum disorder (ASD), ALS, AD, PD, Epilepsy, MS, total body BMD, femoral BMD, and Heel BMD. These GWAS studies do not overlap in sample composition with the UKB cohort used for UDIP-FA in our analysis.

Numerous polygenic profiles of complex traits were generated for each trait using PLINK^76^ based on the output of PRS-CS. To evaluate the association between the polygenic risk scores (PRS) of various traits and UDIP-FA, we applied canonical correlation analysis (CCA), adjusting for covariates used in the GWAS, including age, sex, the first 10 genetic principal components, head size, head position in the scanner, scanner table position, assessment center location, and date of assessment. All *P*-values were derived from *F*-tests and corrected for multiple comparisons using the Bonferroni correction.

### Collection of risk gene for brain disorders

Risk genes associated with brain disorders were compiled from various resources: (1) risk genes of ADHD were sourced from the ADHDgene database (http://adhd.psych.ac.cn), selecting only those with support from at least 60% of all studies included in the database^78^; (2) risk genes of ASD were downloaded from the AutDB database (http://autism.mindspec.org/autdb) and were supplemented with risk genes from recent studies^79,80^; (3) risk genes of SCZ were obtained from the SZGene database (http://www.szgene.org/) and from research by Wang *et al.*^47,81^; (4) risk genes of BIP were gathered from DisGeNet^82^; (5) risk genes of MDD were downloaded from the Polygenic Pathways database (http://www.polygenicpathways.co.uk/depression.htm); (6) risk genes of AD were obtained from the ALzGene database (http://www.alzgene.org)^83^; (7) risk genes of PD were obtained from the PDGene database (http://www.pdgene.org)^84^. (8) risk genes of MS from the DisGeNET (v25.1.1, https://disgenet.com/)^85^.The full list of these risk genes can be found in Table S13.

### Collection of molecular networks and drug interaction networks

Three extensive molecular networks were utilized in this study to investigate the association between UFAGs and brain disorders: (1) Brain-specific TRNs were obtained from Pearl *et al.*^86^, which included 741 transcription factors (TFs) and 11,092 target genes; (2) Brain-active PPI networks were reconstructed by first downloading the global PPI networks from STRING (https://string-db.org), followed by retaining of protein pairs with physical interaction scores over 700, and proteins active in the adult human brain^87^ (expression value > 0), which finally included 8,568 proteins and 114,892 interaction edges (Table S14); (3) Gene co-expression networks in adult brain were generated by first removing lowly expressed genes (expression value < 0.3) and then retaining top 500,000 significant co-expression pairs between genes (*FDR* < 0.01, Pearson correlation test) using gene expression data from adult human brain^87^.

To construct a high-confidence gene–drug target interaction network, we retrieved drug–gene interaction data from the Drug–Gene Interaction Database (DGIdb, version 5.09)^52^. To ensure biological relevance and clinical applicability, we applied the following stringent filtering criteria: (1) only interactions involving approved drugs were retained; (2) drugs annotated as immunotherapies or anti-neoplastic agents were excluded to focus on agents with broader therapeutic profiles; (3) we restricted interactions to those annotated with direct and well-defined mechanisms, such as inhibitor, agonist, antagonist, activator, blocker, or binder; and The resulting gene–drug target network, where edges indicate reliable, mechanism-defined drug–gene interactions suitable for downstream biological and therapeutic analysis(Table S15).

### Network analysis and visualization

To uncover disorder-specific topologies associated with UFAG, subnetworks corresponding to each disorder and UDIP-FA were extracted using the criterion that one side of the edge is a disorder risk gene or UFAG. The maximal connectivity subgraph was then extracted as the disorder-specific topology. To estimate the correlation of two sub-networks, such as a UFAG-associated sub-network and an PD-related sub-network, the Jaccard distance was employed to measure the enrichment between the networks, defined as follows:

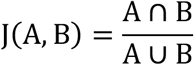

where A was the node set of UFAG-associated sub-network, B was the node set of PD-related sub-network.

For better visualization, we created nodes for each UFAG and other risk genes, connecting all nodes via protein-protein interaction or transcriptional regulation. Finally, we arranged the networks using a perfuse circle layout. In the figure, we only presented the UFAG-associated subnetwork. Network visualization was performed using Cytoscape^88^.

### Correlation analysis between UDIP-FA, UDIP-T1 and UDIP-T2

To evaluate the shared information between 128-dimensional UDIP-FA and UDIPs of other modalities (T1-weight MRI, UDIP-T1; T2-weight MRI, UDIP-T2), we first performed canonical correlation analysis (CCA). Our implementation is based on singular value decomposition (SVD). Given two matrices of different type UDIP, X and Y, we compute their SVD as:

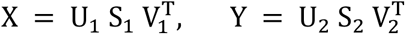

We then calculate the SVD of the inner product 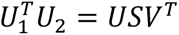 to obtain the canonical correlations S. To quantify how much variance of one space can be explained by the other, we define the variance of X explained by Y and vice versa as follows:

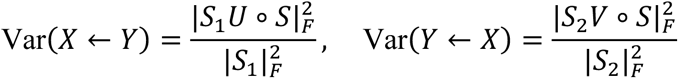

where ∘ denotes element-wise multiplication, and | ⋅ |*_F_* is the Frobenius norm. This formulation reflects the amount of signal preserved across latent representations of different dimensionalities.

To estimate the genetic level association between T1, T2, and FA, we then adopted LDSC^22^ to calculate genetic correlation and using the R language to get the loci overlap.

## Data Availability

All data produced in the present study are available upon reasonable request to the authors

## Data availability

The complete GWAS summary statistics have been deposited in the GWAS Catalog (accession GCP001376). Interactive UDIP-FA visualizations are available at https://udip-fa.github.io/Dim_visualization/.

## Code availability

The codes for this study can be found at Github (https://github.com/ZhiGroup/UDIP-FA)

## Ethics declarations

Our analysis was approved by UTHealth committee for the protection of human subjects under No. HSC-SBMI-20-1323. UKBB has secured informed consent from the participants in the use of their data for approved research projects. UKBB data was accessed via approved project 24247.

## Acknowledgements

This work was partly supported by grants from the National Institute on Aging U01AG070112 and R01AG081398.

## Competing interest

The authors declare that they have no competing interests.

## Notes

### Competing Interest Statement

The authors have declared no competing interest.

### Summary of Updates

This revision updates figures and clarifies text throughout the manuscript. We improved figure quality and readability by increasing resolution, standardizing color scales, refining axis labels and legends, and correcting minor panel annotations. We edited the Results and Methods to clarify data preprocessing, model training, evaluation procedures, and key parameter settings, and we corrected minor numerical inconsistencies. We streamlined language for precision, added or updated citations to situate the work within prior studies, and aligned terminology across sections for consistency. The Abstract and Discussion were revised to reflect these clarifications while avoiding overstatement. Supplementary materials and the data and code availability statements were updated to match the changes in the main text. No new datasets were introduced, the analysis pipeline remains the same, and the principal conclusions are unchanged.

